# Epigenome-wide analyses identify DNA methylation signatures of dementia risk

**DOI:** 10.1101/2020.04.06.20055517

**Authors:** Rosie M. Walker, Mairead L. Bermingham, Kadi Vaher, Stewart W. Morris, Toni-Kim Clarke, Andrew D. Bretherick, Yanni Zeng, Carmen Amador, Konrad Rawlik, Kalyani Pandya, Caroline Hayward, Archie Campbell, David J. Porteous, Andrew M. McIntosh, Riccardo E. Marioni, Kathryn L. Evans

## Abstract

**INTRODUCTION:** Dementia pathogenesis begins years before clinical symptom onset, necessitating the understanding of premorbid risk mechanisms. Here, we investigated potential pathogenic mechanisms by assessing DNA methylation associations with dementia risk factors in Alzheimer’s disease (AD)-free participants.

**METHODS:** Associations between dementia risk measures (family history, genetic risk score (GRS), and dementia risk scores (combining lifestyle, demographic and genetic factors) and whole-blood DNA methylation were assessed in discovery and replication samples (n=∼400 – ∼5,000) from Generation Scotland.

**RESULTS:** AD genetic risk and two risk scores were associated with differential methylation. The GRS predominantly associated with methylation differences in *cis* but also identified a genomic region implicated in Parkinson’s disease. Loci associated with the risk scores were enriched for those previously associated with body mass index and alcohol consumption.

**DISCUSSION:** Dementia risk measures show widespread association with blood-based methylation, which indicates differences in the processes affected by genetic and demographic/lifestyle risk factors.

## 1. Introduction

The pathophysiology of dementia begins many years, possibly decades, before the emergence of clinical symptoms [1]. This long prodromal phase highlights the need for preventative strategies prior to the development of irreversible brain damage. As such, understanding premorbid risk mechanisms is critical. Several approaches to identify individuals at risk of developing dementia have been devised, including the summation of genetic risk, in the form of genetic risk scores (GRSs), the consideration of family history, and the calculation of risk scores, which incorporate multiple lifestyle, demographic and genetic risk factors [2-4].

DNA methylation is an epigenetic modification that is associated with gene expression variation. Altered gene expression has been identified in the blood and post-mortem brains of AD patients [5, 6], and post-mortem brain-based studies have identified associations between DNA methylation and AD and its neuropathological hallmarks [7-9]. Blood-based studies, whilst limited by small sample sizes, have also found evidence for AD-associated methylation differences [10, 11]. It is not, however, possible to determine from these studies whether methylation differences precede AD onset, making them potentially aetiologically informative or whether they reflect ongoing pathology, compensatory mechanisms and/or treatment effects. Studies that have identified associations between variation in blood-based DNA methylation and risk factors for dementia (e.g. carrying the *APOE* ε4 haplotype [12, 13], ageing [14], and obesity [15]) suggest that the assessment of methylation in this tissue may yield insights into the pathways and processes that lead to dementia.

In this study, by assessing associations between multiple measures of dementia risk and blood-based DNA methylation in AD-free participants, we aim to further understanding of the mechanisms conferring dementia risk and characterise the role of methylation in these processes.

## 2. Methods

### 2.1. Participants

Participants were drawn from the Generation Scotland: Scottish Family Health Study (GS:SFHS) [16, 17]. The cohort comprises ∼24,000 participants aged ≥18 years at recruitment. At a baseline clinical appointment, participants were phenotyped for a range of health, demographic and lifestyle factors, and provided physical measurements and samples for DNA extraction. GS:SFHS has been granted ethical approval from the NHS Tayside Committee on Medical Research Ethics, on behalf of the National Health Service (05/S1401/89) and has Research Tissue Bank Status (15/ES/0040).

### 2.2. Calculation of dementia risk scores

Four dementia risk scores, henceforth referred to as CAIDE1, CAIDE2 [2], Li [3] and Reitz [4] were calculated using data that was collected at GS:SFHS enrolment or obtained through record linkage (see Figure 1, Supplementary Methods and Supplementary Table 1 for information on the contributing variables). To generate each risk score, the contributing variables were scaled and weighted according to the original studies and summed. Each score was calculated for participants within the appropriate age-range (CAIDE1/2: 39-64 years [2]; Li: ≥60 years [3]; Reitz: ≥65 years [4]).

**Figure 1.**
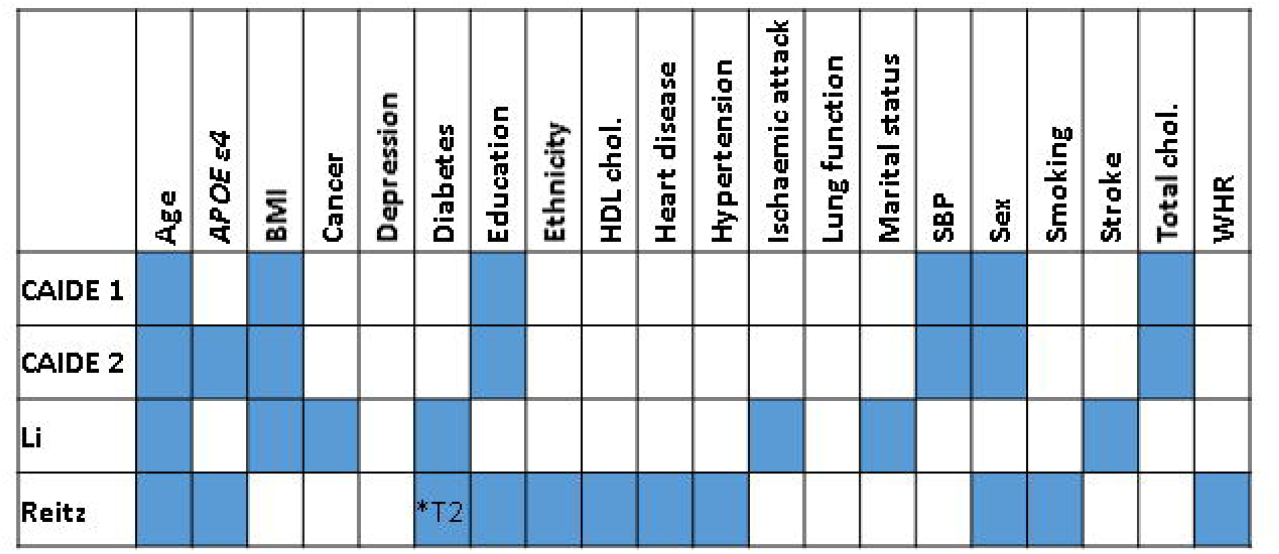
The variables contributing to each dementia risk score are indicated by filled blue boxes. Abbreviations: *APOE* ε4 = Apolipoprotein epsilon 4; BMI = body mass index; HDL = high density lipoprotein; chol = cholesterol; SBP = systolic blood pressure; WHR = waist-hip ratio.

### 2.3. Genotyping and calculation of Alzheimer’s disease genetic risk score

GS:SFHS genotyping has been described previously [18, 19] (Supplementary Methods). The AD GRS was calculated using the lead SNP from each of the 26 genome-wide significant loci identified through a meta-analysis of parental AD and AD [20] (Supplementary Table 2). Each participant’s score was generated by summing their dosage of each risk allele, weighted by the corresponding GWAS effect estimate.

### 2.4. DNA methylation profiling

Whole blood DNA methylation was profiled using the Infinium MethylationEPIC BeadChip (Illumina Inc.) in two sets of GS:SFHS participants at two separate times, leading to a natural discovery (n=5,190) and replication (n=4,588) design, as described previously [21-23] (Supplementary Methods). Discovery and replication sample normalisation was performed separately and the data were converted to M-values. Participants in the replication sample were unrelated (SNP-based relatedness<.05) to each other and/or those in the discovery sample. A correction for relatedness was applied to the discovery sample (Supplementary Methods).

Prior to analyses, poor-performing probes (Supplementary Methods), sex chromosome probes, participants with unreliable self-report data, suspected XXY genotype, or self-reported AD (n=5) were excluded. The final discovery dataset comprised 777,193 loci in 5,087 participants; the replication dataset comprised 773,860 loci in 4,450 participants. Subsequent analyses of the methylation data were carried out using R versions 3.6.0. or 3.6.1.[24].

### 2.5. Epigenome-wide association studies

EWASs were performed using linear regression modelling, implemented in limma [25]. CpG sites (M-values) were modelled as the dependent variable and the dementia risk measure was the predictor-of-interest. Additional covariates included in the standard models are detailed below:

#### Discovery sample

CpG site (M-values pre-corrected for relatedness, estimated cell count proportions and processing batch) ∼ dementia risk measure + age + sex + smoking status + pack years + 20 methylation principal components

#### Replication sample

CpG site (M-values) ∼ dementia risk measure+ age + sex + smoking status + pack years + estimated cell count proportions (granulocytes, natural killer cells, B-lymphocytes, CD4+ T-lymphocytes and CD8+ T-lymphocytes) + processing batch + 20 methylation principal components

The variables “Smoking status”, “pack years” and the methylation principal components are explained in the Supplementary Methods.

A number of sensitivity analyses for the CAIDE1 score were performed in which additional covariates were included one-by-one, using the same thresholds for categorising continuous variables as implemented in the risk score. These were: body mass index (BMI; ≤30kg/m^2^ or >30 kg/m^2^); systolic blood pressure (SBP; ≤140 mm Hg or >140 mm Hg); total cholesterol (TC; ≤6.5 mmol/L or > 6.5 mmol/L); years in education (≥10, >6 and <10, or ≤6); self-reported alcohol consumption (log_10_-transformed (+1) units of alcohol/week), and a DNA methylation alcohol consumption score derived using the R package dnamalci [26, 27].

Limma was used to calculate empirical Bayes moderated t-statistics from which *P* values were obtained. The significance threshold in the discovery sample was *P*≤3.6 × 10^−8^ [28]. Sites attaining significance in the discovery sample were assessed in the replication sample using a Bonferroni-adjusted threshold of 0.05/no. sites assessed.

### 2.6. EWAS meta-analysis

Inverse standard error-weighted fixed effects meta-analyses of the discovery and replication EWAS results were performed using METAL [29]. Sites attaining a meta-analysis *P*≤3.6 × 10^−8^ were considered significant.

### 2.7. Identification of differentially methylated regions

Differentially methylated regions (DMRs) were identified using the dmrff.meta function in dmrff [30]. DMRs were defined as regions containing 2-30 sites with consistent direction of effect and EWAS meta-analysis *P* values ≤.05 separated by ≤500 bp. DMRs with Bonferroni-adjusted *P* values ≤.05 were declared significant.

### 2.8. EWAS and GWAS catalog look-ups

The GWAS Catalog v1.0.2. was downloaded from https://www.ebi.ac.uk/gwas/docs/file-downloads (16/12/19) and queried using gene names annotated to probes containing meta-DMPs for the phenotype-of-interest. GWAS traits enriched for association (*P*≤1 × 10^−5^) with genes containing meta-DMPs were identified using Fisher’s exact test. Enrichment was declared significant when *P*≤1.26 × 10^−5^ (0.05/3980 traits assessed).

The EWAS Catalog was downloaded from http://www.ewascatalog.org/ (03/07/19) and queried using the significant DMPs’ probe IDs. EWAS traits enriched for association (*P*≤1 × 10^−5^) with meta-DMPs were identified using Fisher’s exact test. Enrichment was declared significant when *P*≤3.31 × 10^−4^ (0.05/151 traits assessed).

### 2.9. Gene ontology/KEGG pathway analyses

Gene ontology (GO) and KEGG pathway analyses were implemented using a modified version of the missMethyl gometh function [31] (Supplementary Methods). The target list comprised probes showing suggestive association with the phenotype-of-interest (*P*≤1 × 10^−5^) in the EWAS or DMR analysis and the gene universe included all probes in the analyses. Enrichment was assessed using a hypergeometric test, accounting for the bias arising from the variation in the number of probes per gene. Significance thresholds of *P*≤2.20 × 10^−6^ and *P*≤1.48 × 10^−4^ were applied to allow for a Bonferroni-correction for the 22,750 GO terms and 337 KEGG pathways assessed, respectively.

### 2.10. Identification of meQTLs

Methylation quantitative trait loci for the AD GRS-associated DMPs were identified using the discovery sample (Bretherick et al., in preparation). Following quality control (described in 2.4), the data was normalised and corrected as described previously [32] (Supplementary Methods). The resulting residuals were inverse rank normal transformed and entered as the dependent variable in simple linear model GWASs to identify meQTLs. GWASs were implemented using REGSCAN v0.5 [33].

SNPs that were associated with a DMP with *P*≤5 × 10^−8^/49 (Bonferroni correction for the 49 DMPs for which meQTL results were available) and had a MAF>0.01 were declared to be meQTLs.

## 3. Results

### 3.1. EWAS sample demographics

Participant numbers and sample demographic information are shown in Supplementary Table 3.

### 3.2. Genetic risk for Alzheimer’s disease

#### 3.2.1. Identification of differentially methylated positions

An EWAS of the AD GRS identified 32 DMPs in the discovery sample (1.06 × 10^−30^ ≤*P*≤ 2.22 × 10^−8^; Supplementary Table 4). Of these, 31 showed replicated association (1.07 × 10^−30^ ≤*P*≤ 8.33 × 10^−4^; Supplementary Table 5). Meta-analysis of the discovery and replication samples identified 68 DMPs (6.15 × 10^−48^ ≤*P*≤ 3.45 × 10^−8^; Table 1; Supplementary Table 6; Figure 2).

**Table 1.** Top 20 DMPs associated with the AD GRS in a meta-analysis of the discovery and replication samples.

| ID | Chr. | BP* | Gene Symbol | Effect | SE | P value |
| --- | --- | --- | --- | --- | --- | --- |
| cg10757760 | 2 | 127893054 | | 0.0568 | 0.0039 | $6.15 \times 10^{-48}$ |
| cg04441687 | 11 | 85869322 | | 0.0586 | 0.0041 | $1.12 \times 10^{-46}$ |
| cg26631131 | 19 | 45240591 | | 0.0385 | 0.0029 | $1.56 \times 10^{-39}$ |
| cg02887598 | 2 | 127841945 | *BIN1* | -0.0986 | 0.0085 | $2.51 \times 10^{-31}$ |
| cg19116668 | 7 | 99932089 | *PMS2L1* | 0.049 | 0.0051 | $1.54 \times 10^{-21}$ |
| cg18959616 | 11 | 85814918 | | 0.0656 | 0.0072 | $5.85 \times 10^{-20}$ |
| cg02521229 | 11 | 60019236 | | 0.0658 | 0.0073 | $1.61 \times 10^{-19}$ |
| cg03579757 | 7 | 100091793 | *NYAP1* | 0.0342 | 0.0038 | $2.33 \times 10^{-19}$ |
| cg16618979 | 7 | 143108841 | *AC092214.10* | 0.0974 | 0.0109 | $5.11 \times 10^{-19}$ |
| cg03526776 | 6 | 41159608 | *TREML2* | -0.0402 | 0.0045 | $7.06 \times 10^{-19}$ |
| cg11461311 | 2 | 127782614 | *RP11-521O16.1;RP11-521O16.2* | 0.0321 | 0.0036 | $1.31 \times 10^{-18}$ |
| cg00436254 | 2 | 127862614 | *BIN1* | 0.0256 | 0.003 | $6.76 \times 10^{-18}$ |
| cg06750524 | 19 | 45409955 | *APOE* | 0.05 | 0.0059 | $2.18 \times 10^{-17}$ |
| cg23423086 | 11 | 85856245 | | -0.0365 | 0.0043 | $2.37 \times 10^{-17}$ |
| cg22906224 | 7 | 99728672 | *AC073842.19* | -0.0392 | 0.0047 | $3.55 \times 10^{-17}$ |
| cg05908241 | 7 | 143109367 | *AC092214.10* | 0.0283 | 0.0035 | $6.62 \times 10^{-16}$ |
| cg17830204 | 7 | 99819110 | *GATS;PVRIG;STAG3;AC005071.1* | 0.0308 | 0.0039 | $4.78 \times 10^{-15}$ |
| cg19590598 | 2 | 127782813 | *RP11-521O16.1;RP11-521O16.2* | 0.0282 | 0.0036 | $5.22 \times 10^{-15}$ |
| cg08871934 | 10 | 11720283 | | -0.0343 | 0.0044 | $9.59 \times 10^{-15}$ |
| cg09555818 | 19 | 45449301 | *APOC2;APOC4* | -0.0498 | 0.0065 | $1.24 \times 10^{-14}$ |

Abbreviations: BP, base position; Chr., chromosome; SE, standard error
\*Base position in genome assembly hg19/GRCh37

**Figure 2.**
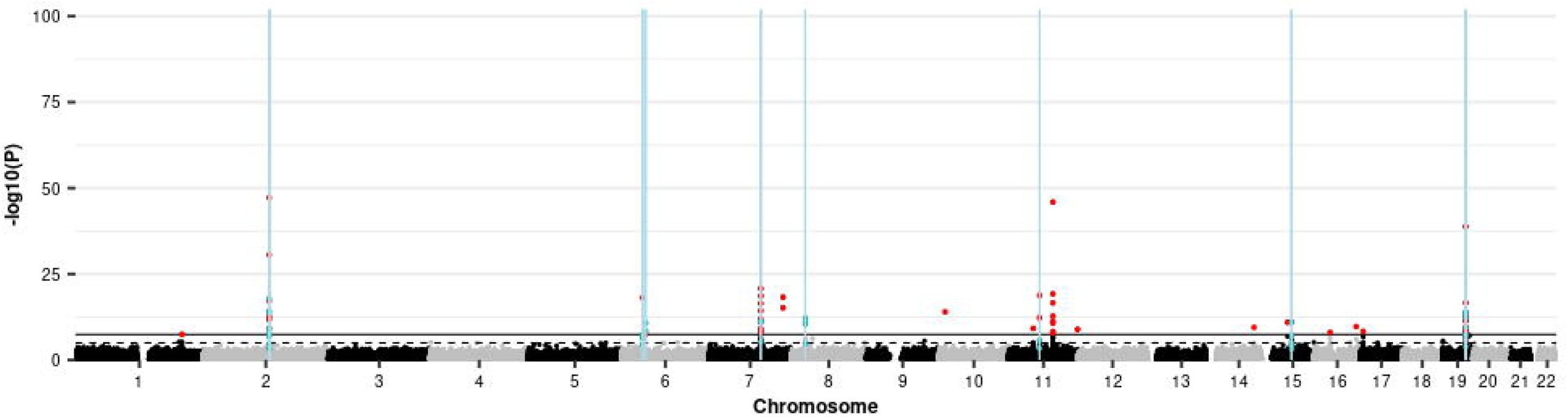
Manhattan plot showing the results of the EWAS meta-analysis of the AD GRS and the positions of DMRs identified in a meta-DMR analysis. Each point represents one of the 772,453 loci included in the EWAS meta-analysis, with the point’s position being determined by genomic position (x-axis) and significance in the EWAS meta-analysis (–log_10_ *P* value; y-axis). Sites attaining genome-wide significance (*P* ≤ 3.6 × 10^−8^) are indicated in red and those that are involved in a significant DMR (Bonferroni-correct *P* ≤ 0.05) are indicated in blue. The locations of DMRs are further indicated by vertical blue lines. The solid horizontal line is the threshold for genome-wide significance (*P* ≤ 3.6 × 10^−8^) and the dashed line indicates a suggestive significance threshold (*P* ≤ 1 × 10^−5^).

Sixty-one of the 68 meta-DMPs were located within 18 of the 26 GWAS loci used to produce the GRS [20]. meQTLs were identified for 49/68 DMPs: of these, 48 were associated in *cis* with genetic variants located in one the GWAS loci [20]. None of these 48 DMPs were associated in *trans* with genetic variation in other GWAS loci. Methylation at the remaining DMP, cg14354618, was associated in *trans* with genetic variation on chromosome 19 (chr19: 868083-1188756; hg19/GRCh37), which overlaps a GWAS locus [20].

Querying the GWAS catalog [34] with the 34 gene names annotated to the 68 meta-DMPs unsurprisingly identified many terms related to AD and its neuropathological hallmarks (Supplementary Table 7) (the most significant being “Alzheimer’s disease or family history of Alzheimer’s disease” (*P*=1.77 × 10^−27^)). No significant enrichment was identified when querying the EWAS catalog; however, this catalogue comprises results from studies using the 450K array on which only 30/68 meta-DMPs were measured.

#### 3.2.2. Identification of differentially methylated regions

The DMR meta-analysis identified 18 significant DMRs comprising 41 CpGs, of which 20 were identified by the meta-EWAS (Supplementary Table 8; Figure 2). Seventeen of the DMRs overlap with loci that were in the GWAS [20], and the eighteenth is located <7kb from the nearest GWAS locus. The longest DMR spans a 302 bp region ∼13.7 kb upstream of *BIN1*, whilst the most significant spans a 199 bp region ∼22.8 kb downstream of *BIN1*. Both show increased methylation with increased GRS.

GO analysis using the combined DMP and DMR results identified 18 terms, the most significant of which was “amyloid-beta formation” (*P*=3.68 × 10^−10^; Table 2). No significant KEGG pathways were identified.

**Table 2.**
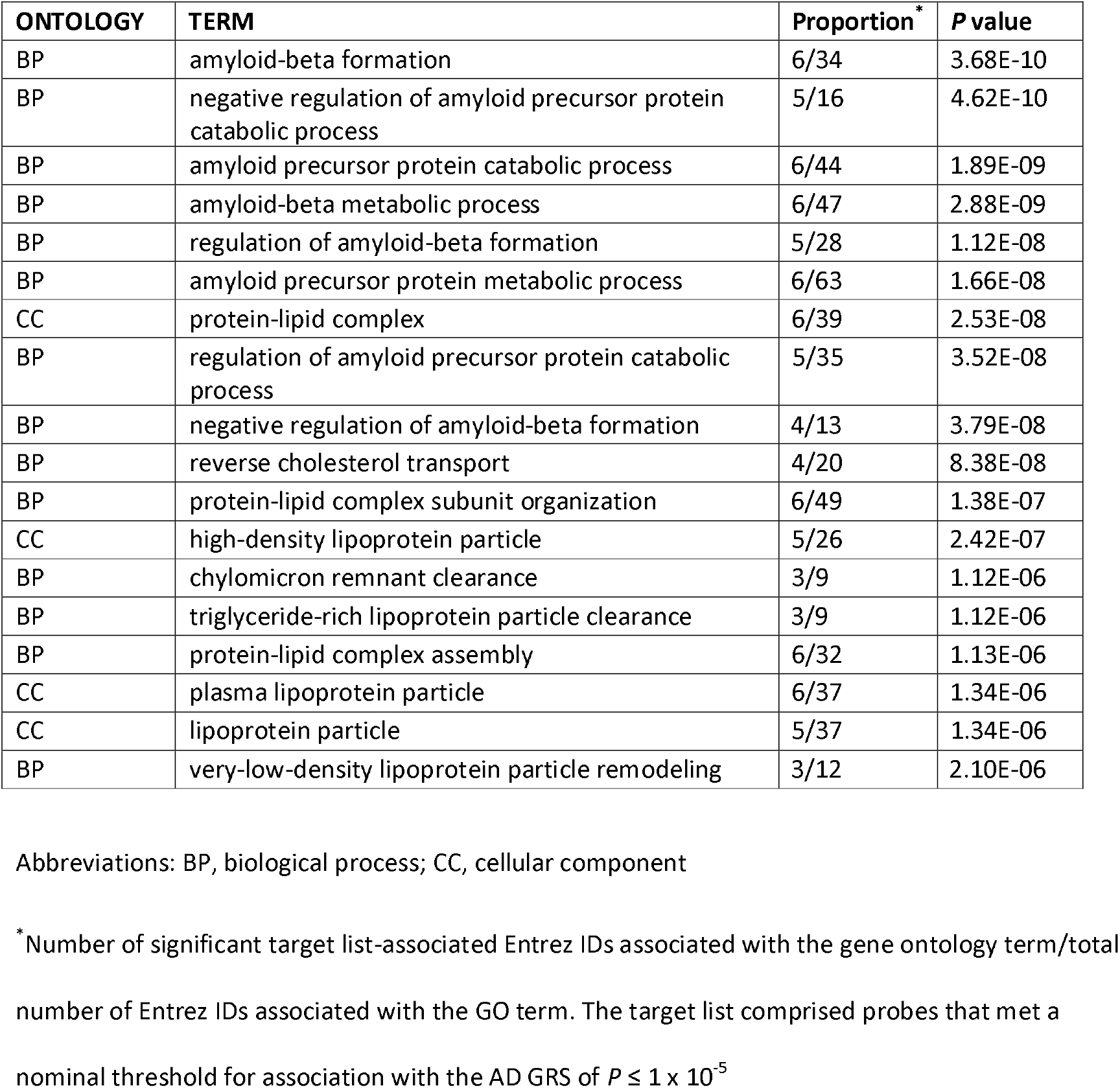
GO terms showing significant enrichment for probes where methylation is associated the AD GRS

### 3.3. Mid-life dementia risk scores

The CAIDE1 and CAIDE2 risk scores assesses the risk of developing dementia in 20 years’ time in individuals aged 39-64 years [2]. CAIDE2 takes into account the same risk factors as CAIDE1 (with different weightings) and also considers *APOE* ε4 carrier status.

#### 3.3.1. Identification of differentially methylated positions

An EWAS of the CAIDE1 score in the discovery sample identified 76 DMPs (3.29 × 10^−20^ ≤*P*≤ 3.49 × 10^−8^; Supplementary Table 9), of which 65 replicated (7.76 × 10^−18^ ≤*P*≤ 6.48 × 10^−4^; Supplementary Table 10). Meta-analysis of the discovery and replication samples identified 227 DMPs (1.20 × 10^−29^ ≤*P*≤ 3.58 × 10^−8^; Figure 3; Table 3; Supplementary Table 11).

**Table 3.** Top 20 DMPs associated with the CAIDE1 risk score in a meta-analysis of the discovery and replication samples.

| ID | Chr. | BP | Gene Symbol | Direction | Effect | SE | P value |
| --- | --- | --- | --- | --- | --- | --- | --- |
| cg06690548 | 4 | 139162808 | *SLC7A11* | -- | -0.0766 | 0.0068 | 1.20E-29 |
| cg19758958 | 11 | 62319222 | *AHNAK* | -- | -0.0256 | 0.0024 | 4.22E-27 |
| cg11024682 | 17 | 17730094 | *SREBF1* | ++ | 0.026 | 0.0025 | 5.31E-26 |
| cg14476101 | 1 | 120255992 | *PHGDH* | -- | -0.0391 | 0.0039 | 1.14E-23 |
| cg00574958 | 11 | 68607622 | *CPT1A* | -- | -0.0413 | 0.0043 | 8.30E-22 |
| cg06500161 | 21 | 43656587 | *ABCG1* | ++ | 0.0225 | 0.0024 | 2.10E-21 |
| cg19693031 | 1 | 145441552 | *TXNIP* | -- | -0.0369 | 0.004 | 8.74E-21 |
| cg22699725 | 1 | 207242586 | *PFKFB2* | ++ | 0.0275 | 0.003 | 6.89E-20 |
| cg00683922 | 1 | 207242569 | *PFKFB2* | ++ | 0.0272 | 0.003 | 2.70E-19 |
| cg05325763 | 11 | 68607719 | *CPT1A* | -- | -0.0394 | 0.0044 | 2.90E-19 |
| cg22976567 | 1 | 156074182 | *LMNA* | -- | -0.026 | 0.0029 | 7.06E-19 |
| cg02715788 | 8 | 119974400 |  | -- | -0.0225 | 0.0026 | 4.44E-18 |
| cg18120259 | 6 | 43894639 | *LOC100132354* | -- | -0.0209 | 0.0024 | 4.49E-18 |
| cg11376147 | 11 | 57261198 | *SLC43A1* | -- | -0.0236 | 0.0028 | 2.98E-17 |
| cg00163198 | 11 | 130767760 | *SNX19* | ++ | 0.0237 | 0.0028 | 4.89E-17 |
| cg16246545 | 1 | 120255941 | *PHGDH* | -- | -0.0255 | 0.003 | 5.16E-17 |
| cg01270753 | 9 | 101944336 | *RP11-96L7.2* | -- | -0.034 | 0.0041 | 5.27E-17 |
| cg08857797 | 17 | 40927699 | *VPS25* | ++ | 0.0236 | 0.0028 | 5.86E-17 |
| cg16740586 | 21 | 43655919 | *ABCG1* | ++ | 0.0243 | 0.0029 | 7.52E-17 |
| cg26457483 | 1 | 120256112 | *PHGDH* | -- | -0.0308 | 0.0037 | 1.76E-16 |

Abbreviations: BP, base position; Chr., chromosome; SE, standard error
\*Base position in genome assembly hg19/GRCh37

**Figure 3.**
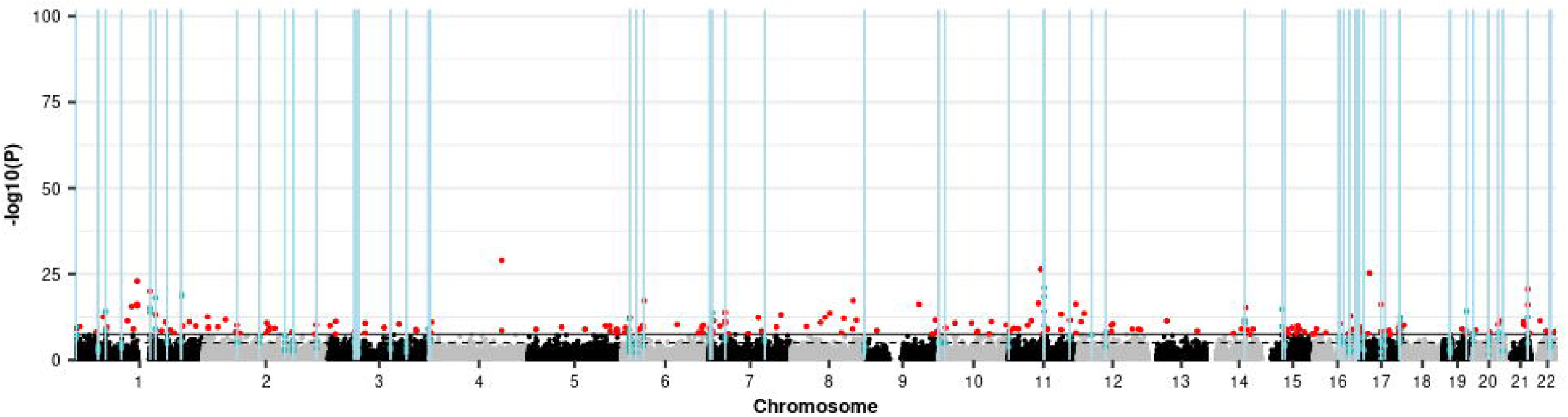
Manhattan plot showing the results of the EWAS meta-analysis of the CAIDE1 dementia risk score and the positions of DMRs identified in a meta-DMR analysis. Each point represents one of the 772,453 loci included in the EWAS meta-analysis, with the point’s position being determined by genomic position (x-axis) and significance in the EWAS meta-analysis (–log_10_ *P* value; y-axis). Sites attaining genome-wide significance (*P* ≤ 3.6 × 10^−8^) are indicated in red and those that are involved in a significant DMR (Bonferroni-correct *P* ≤ 0.05) are indicated in blue. The locations of DMRs are further indicated by vertical blue lines. The solid horizontal line is the threshold for genome-wide significance (*P* ≤ 3.6 × 10^−8^) and the dashed line indicates a suggestive significance threshold (*P* ≤ 1 × 10^−5^).

An EWAS of the CAIDE2 score in the discovery sample identified 18 DMPs (1.96 × 10^−17^ ≤*P*≤ 3.24 × 10^−8^; Supplementary Table 12), of which 17 replicated (4.91 × 10^−12^ ≤*P*≤ 2.01 × 10^−3^; Supplementary Table 13). Meta-analysis of the discovery and replication samples identified 59 DMPs (3.56 × 10^−22^ ≤*P*≤ 3.21 × 10^−8^; Supplementary Table 14). Fifty-four of the CAIDE2 meta-DMPs were also identified by the CAIDE1 meta-analysis; given this overlap, subsequent analyses focus on CAIDE1.

The CAIDE1-associated meta-DMPs were explored using the EWAS and GWAS catalogs [34]. Significant enrichment was identified for 16 EWAS traits/conditions, with “Body mass index” being the most significantly enriched (*P*=2.96 × 10^−118^; Supplementary Table 15). Two alcohol-related traits: “alcohol consumption per day” (*P*=9.70 × 10^−29^) and “gamma-glutamyl transferase” (*P*=8.55 × 10^−25^; Table 6) also showed enrichment. GWAS catalog enrichment analysis identified only one significant term, “Eosinophil counts” (*P*=4.97 × 10^−8^).

##### 3.3.1.1. Sensitivity analyses

The extent to which the BMI component of the CAIDE1 score drives the observed CAIDE1 associations was assessed by performing an EWAS meta-analysis in which BMI was included as an additional covariate. Co-varying for BMI resulted in only 11 of the original 227 meta-DMPs remaining significant (Supplementary Table 11), with the correlation between the effect estimates for all sites between the two analyses being *r*=0.745 (95% CI=0.744-0.746). Compared with BMI, co-varying for other components of the CAIDE1 score resulted in a larger numbers of CAIDE1 meta-DMPs remaining significant (SBP: 94/227; TC: 190/227; education: 221/227; Supplementary Table 11); correlations with the effect estimates of the original analysis were higher (SBP: *r*=0.815, 95% CI=0.815-0.816; TC: *r*=0.938, 95% CI=0.937-0.938); education: *r*=0.990, 95% CI=0.990-0.990).

##### 3.3.1.2. Assessment of the involvement of alcohol consumption in the CAIDE1 EWAS results

Twenty-four of the 88 meta-DMPs that are represented on the 450K array, including the most significant DMP, cg06690548, have previously been associated with alcohol consumption (*P*≤1 × 10^−5^) [26]. Similarly, an EWAS meta-analysis of self-reported alcohol consumption in our methylation sample identified 5599 DMPs at a suggestive threshold of *P*≤1 × 10^−5^ (Clarke et al., in preparation); of these, 49 show a significant association with CAIDE1 with a consistent direction of effect. This overlap is highly significant (*p*<2 × 10^−16^).

As alcohol consumption showed a small but significant correlation with CAIDE1 score (*r*=0.091; 95% CI=0.065-0.118; *P*=2.60 × 10^−11^), the potential for alcohol consumption to drive the observed associations between CAIDE1 and DNA methylation was assessed by including alcohol consumption measured by (i) self-report or (ii) a polyepigenetic risk score [26, 27] as an additional covariate in the CAIDE1 EWAS. Neither measure of alcohol consumption resulted in a substantial change in effect estimates (self-reported: *r* = 0.948, 95% CI = 0.948-0.948; DNA methylation score: *r* = 0.993, 95% CI = 0.993-0.993), with 166 and 191 of the 227 CAIDE1 meta-DMPs remaining significant after the inclusion of the self-reported and DNA methylation score coefficients, respectively (Supplementary Table 11).

#### 3.3.2. Identification of differentially methylated regions

The DMR meta-analysis of the discovery and replication samples identified 57 CAIDE1-associated DMRs (all Bonferroni-adjusted *P*<0.044; Supplementary Table 16) each comprising two to seven CpGs. In total, the 57 DMRs involve 179 sites, of which 35 were significant in the EWAS meta-analysis. The most significant DMR (Bonferroni-adjusted *P*=1.67 × 10^−20^) comprises four hypomethylated CpGs spanning a 115 bp intronic region of *CPT1A* (chr11: 68607622-68607737; hg19/GRCh37). The longest DMR spans a 1.1 kb intronic region of *JARID2* (chr6: 15504844-15505949; hg19/GRCh37).

GO and KEGG pathway analyses found no enrichment of biological processes or pathways amongst the CAIDE1 DMP or DMR CpG sites (min. *P*_GO_=8.46 × 10^−4^; min. P_KEGG_=3.51 × 10^−3^).

### 3.4. Other measures of dementia risk

The other measures of dementia risk assessed were (i) dementia FH and (ii) two late-life dementia risk scores that predict the risk of developing dementia in those aged over 60 or 65 [3, 4]. EWASs of the discovery sample (min. *P*_FH_=8.47 × 10^−7^; min. *P*_Li_=3.91 × 10^−8^; min *P*_Reitz_=1.58 × 10^−6^), meta-EWASs of the discovery and replication samples (min. *P*_FH_=1.15 × 10^−6^; min. *P*_Li_=1.80 × 10^−6^; min *P*_Reitz_=8.99 × 10^−7^), and DMR analyses (min. Bonferroni-adjusted *P*_FH_=0.439; min. Bonferroni-adjusted *P*_Li_=0.208; min Bonferroni-adjusted *P*_*Reitz*_=1) failed to identify any significant associations.

## 4. Discussion

We have assessed DNA methylation associations with a range of dementia risk measures in large discovery and replication samples comprising AD-free participants and report multiple loci as being associated with AD genetic risk and two multifactorial mid-life risk scores.

All but one of the loci associated with the AD GRS were located within 30 kb of the GWAS loci used to derive the GRS [20], with meQTL analysis supporting involvement of *cis* meQTLs. Only one DMP, cg14354618 on chromosome 11, was an exception to this pattern, being associated with *trans* meQTLs in a GWAS risk locus on chromosome 19. cg14354618 is located in a CpG island in *AP001979*.*1*. Genetic variation annotated to *AP001979*.1 has been associated with Parkinson’s disease [35, 36], body fat percentage [37], and sugar consumption [38] but has not been associated with AD in large-scale GWASs [39, 40]. There is a degree of overlap between the clinical features and pathologies associated with AD and Parksinon’s disease, with certain genetic variants being associated with both [41, 42]. Moreover, obesity and hyperglycaemia have been implicated as dementia risk factors [43]. Taken together, these findings suggest this locus to be a plausible AD-risk locus, which warrants further investigation.

Considering both the meta-DMP and meta-DMR results, two regions harbour a large number of AD GRS-associated sites. These regions contain (i) *BIN1* and (ii) *PVRL2, APOE, APOC4* and *APOC2* (henceforth referred to as the *APOE* locus). The *APOE* locus has not previously been identified by brain-based EWASs of AD neuropathological hallmarks [7, 44] or a blood-based AD case-control EWAS [44]; larger samples might be required to detect association between methylation at this locus and AD.

In contrast, several studies have identified altered methylation of *BIN1* in AD patients or in association with AD neuropathological hallmarks [7, 45, 46]. These findings are of particular interest as altered *BIN1* brain expression has been reported in AD [47-49] and DNA methylation has been suggested to regulate *BIN1*’s expression [50]. We identified a mixture of hyper- and hypomethylation in the upstream region and gene body and hypermethylation in the downstream region. Whilst none of the identified sites directly replicated those identified by previous studies, it is noteworthy that one of our hypermethylated meta-DMPs (cg18813565) is located only 31bp from a site (that failed our quality control) at which increased methylation in the dorsolateral prefrontal cortex has been associated with neuritic plaque burden and AD diagnosis [45]. Moreover, this site contributes to a hypermethylated DMR that spans a 199bp region located ∼23kb downstream of *BIN1*. This region overlaps with non-coding RNAs, RP11-521O16.1 and RP11-521O16.2, suggesting the possibility that altered methylation of these non-coding RNAs might alter *BIN1* expression. This hypothesis should be assessed by future studies.

The CAIDE1 risk score is a composite score formed by the weighted summation of age, sex, BMI, years in education, systolic blood pressure and total cholesterol [2]. It is designed for the prediction of dementia in 20 years’ time in individuals aged 39-64 years, which it does with an AUC of 0.77 (95% CI 0.71–0.83). Since age and sex were covariates in our analytical models, the differential methylation observed in this study reflects the modifiable “lifestyle” components of the score. Our analyses revealed BMI to be the primary driver of the CAIDE1-associated methylation differences. We identified significant overlap between the sites associated with CAIDE1 and those that have previously been associated with alcohol consumption. Strikingly, the most significant CpG in our analysis of CAIDE1 was also the most significant CpG in a recent GS:SFHS alcohol consumption EWAS (Clarke et al., in preparation). Whilst alcohol consumption was significantly correlated with CAIDE1 score, it could not account for the CAIDE1-associated differences in methylation. This finding is of interest in light of the observed associations between excessive alcohol consumption and dementia risk [51]. Our findings suggest that the risk factors contributing to the CAIDE1 score and alcohol consumption might confer risk for AD via independent effects on common pathways.

The loci implicated by our analyses of the AD GRS and the CAIDE1 score did not overlap, nor did they implicate common genes. This finding is intriguing as it suggests that lifestyle and genetic risk factors for dementia may act via separate pathways.

We did not observe any DNA methylation associations with AD FH or two late-life dementia risk scores. The lack of associations with AD FH is somewhat surprising as this has previously been shown to be a good AD proxy-phenotype [20]. As AD FH was self-reported, inaccurate reporting may have affected our results. Our failure to observe significant associations for these traits may also reflect a lack of statistical power, particularly as the samples available for the late-life dementia risk scores were relatively small.

It is important to note some additional strengths and limitations to our study. Whilst it would clearly be desirable to study DNA methylation in brain tissue, growing evidence highlights the contribution of systemic factors to dementia pathogenesis [52]. Thus, methylation studies in the blood are necessary to provide a holistic characterisation of the processes that contribute to dementia development. Moreover, profiling blood methylation permits both longitudinal analyses to characterise the dynamic processes underlying dementia pathogenesis and biomarker identification. Other limitations concern the quality of the variables used in the dementia risk scores: self-reporting may have resulted in errors, and the blood samples used for cholesterol measurements were not taken at a consistent time of day or after a consistent fasting length.

Here we present the first comprehensive characterisation of associations between blood DNA methylation and dementia risk, performed in the largest single-cohort methylation samples collected to-date. We identify several CpGs where methylation is associated with dementia risk measures, identify a putative novel AD risk locus, and find genetic and lifestyle risk factors to be associated with methylation levels at different CpGs. Our findings suggest a number of hypotheses for assessment by future studies, which should include longitudinal assessments of the causal nature of methylation in dementia pathogenesis.

## Data Availability

According to the terms of consent for GS:SFHS, access to data must be reviewed by the GS Access Committee.

## Acknowledgements

This work was supported by a Wellcome Trust Strategic Award “STratifying Resilience and Depression Longitudinally” (STRADL) [104036/Z/14/Z] to AMM, KLE, CSH, DJP and others, and an MRC Mental Health Data Pathfinder Grant [MC_PC_17209] to AMM and DJP. REM is supported by an Alzheimer’s Research UK major project grant [ARUK-PG2017B-10]. KV is funded by the Wellcome Trust Translational Neuroscience PhD Programme at the University of Edinburgh [108890/Z/15/Z]. ADB would like to acknowledge funding from the Wellcome PhD training fellowship for clinicians [204979/Z/16/Z], the Edinburgh Clinical Academic Track (ECAT) programme. Generation Scotland received core support from the Chief Scientist Office of the Scottish Government Health Directorates [CZD/16/6] and the Scottish Funding Council [HR03006]. Genotyping of the GS:SFHS samples was carried out by the Genetics Core Laboratory at the Clinical Research Facility, Edinburgh, Scotland and was funded by the UK’s Medical Research Council and the Wellcome Trust [104036/Z/14/Z]. DNA methylation profiling of the GS:SFHS samples was funded by the Wellcome Trust Strategic Award [10436/Z/14/Z] with additional funding from a 2018 NARSAD Young Investigator Grant from the Brain & Behavior Research Foundation [27404].

We are grateful to all the families who took part, the general practitioners and the Scottish School of Primary Care for their help in recruiting them, and the whole Generation Scotland team, which includes interviewers, computer and laboratory technicians, clerical workers, research scientists, volunteers, managers, receptionists, healthcare assistants and nurses.

## Supplementary material legends

**Supplementary Table 1**. Details of how the variables contributing to the dementia risk calculators were measured/ascertained in GS: SFHS.

**Supplementary Table 2**. For each SNP contributing to the AD GRS, its chromosomal location (CHR and BP; hg19/GRCh37) are shown, together with the allele assessed (A1) and effect estimate (BETA) from Marioni et al. (2018).

**Supplementary Table 3**. Demographic information for the discovery and replication samples broken down by each measure of dementia risk studied. For each risk measures, the following information is shown: mean age; percentage of female participants; percentages of current smokers, former smokers who quit less than a year ago, former smokers who quit a year or more ago, never smokers and those that didn’t declare their smoking status; mean pack years (amongst current and former smokers). Standard deviations (SD) are shown for all means.

**Supplementary Table 4**. Differentially methylated positions (DMPs) associated (*P* ≤ 3.6 × 10^−8^) with the AD GRS in the discovery sample (n = 5010). For each DMP, the symbol of the gene in which the probe maps (if any), probe location (CHR and MAPINFO; hg19/GRCh37), location relative to genes (FEATURE; Body: within the gene body; TSS200: within 200 bp of the transcription start site; TSS1500: within 1500 bp of the transcription start site) and CpG islands (CpGISLAND), estimated effect (Beta), standard error of the estimated effect (Beta SE), t-statistic and *p*-value are shown.

**Supplementary Table 5**. Differentially methylated positions (DMPs) showing replicated association with the AD GRS. DMPs identified in the discovery sample (n = 5010; *P* ≤ 3.6 × 10^−8^) that showed association (*P* ≤ 0.05/32) with a consistent direction of effect in the replication sample (n = 4380) were deemed to have replicated. The probe ID, symbol of the gene in which the probe maps (if any), probe location (CHR and MAPINFO; hg19/GRCh37), genomic features associated with the probe (Body: within the gene body; TSS200: within 200 bp of the transcription start site; TSS1500: within 1500 bp of the transcription start site), beta coefficient, standard error of the beta, t-statistic and *p*-value in the discovery (dis) and replication (rep) samples are show for each replicated DMP.

**Supplementary Table 6**. Differentially methylated positions (DMPs) associated (*P* ≤ 3.6 × 10^−8^) with the AD GRS in an inverse standard error-weighted fixed effects meta-analysis of the discovery and replication samples (total n = 9390). For each DMP, the probe location (CHR and MAPINFO; hg19/GRCh37), symbol of the gene in which the probe maps (if any), direction of effect in the discovery and replication samples, estimated effect (Beta), standard error of the estimated effect (SE), *p*-value, coordinates of the nearest GWAS locus identified by Marioni et al. (2018), and the distance of the DMP from the GWAS locus are shown.

**Supplementary Table 7**. Traits/conditions in the GWAS catalog showing significant enrichment for the genes harbouring AD GRS-associated meta-DMPs. For each trait or condition, the number of DMP-harbouring genes that have been suggestively (*P*≤1 × 10-5) associated with the phenotype (sig_freq), the number of non-significant CpG-harbouring genes have been suggestively (*P*≤1 × 10-5) associated with the phenotype, and the significance (*P* value) and odds ratio (OR) of the enrichment are shown.

**Supplementary Table 8**. DMRs associated with the AD GRS in a meta-analysis of the discovery and replication samples (total n = 9390). For each DMR, the location (Chr., Start and End, hg19/GRCh37), length, effect size (effect) and standard error of the effect (SE), Bonferroni-adjusted *p*-value (Adj. p-value), number of CpGs in the DMR (N), the identities of these CpGs (CpGs), and the coordinates of the nearest GWAS locus identified by Marioni et al. (2018) are shown.

**Supplementary Table 9**. Differentially methylated positions (DMPs) associated (*P* ≤ 3.6 × 10^−8^) with the CAIDE1 dementia risk score in the discovery sample (n = 3077). For each DMP, the symbol of the gene in which the probe maps (if any), probe location (CHR and MAPINFO; hg19/GRCh37), location relative to genes (FEATURE; Body: within the gene body; TSS200: within 200 bp of the transcription start site; TSS1500: within 1500 bp of the transcription start site) and CpG islands (CpGISLAND), estimated effect (Beta), standard error of the estimated effect (Beta SE), t-statistic and *p*-value are shown.

**Supplementary Table 10**. Differentially methylated positions (DMPs) showing replicated association with the CAIDE1 dementia risk score. DMPs identified in the discovery sample (n = 3077; *P* ≤ 3.6 × 10^−8^) that showed association (*P* ≤ 0.05/76) with a consistent direction of effect in the replication sample (n = 2802) were deemed to have replicated. The probe ID, symbol of the gene in which the probe maps (if any), probe location (CHR and MAPINFO; hg19/GRCh37), genomic features associated with the probe (Body: within the gene body; TSS200: within 200 bp of the transcription start site; TSS1500: within 1500 bp of the transcription start site), beta coefficient, standard error of the beta, t-statistic and p-value in the discovery (dis) and replication (rep) samples are show for each replicated DMP.

**Supplementary Table 11**. Differentially methylated positions (DMPs) associated (*P* ≤ 3.6 × 10^−8^) with the CAIDE1 dementia risk score in an inverse standard error-weighted fixed effects meta-analysis of the discovery and replication samples (total n = 5879). For each DMP, the probe location (CHR and MAPINFO; hg19/GRCh37), symbol of the gene in which the probe maps (if any), direction of effect in the discovery and replication samples, estimated effect (Beta), standard error of the estimated effect (SE) and *p*-value are shown. Results are also shown for the analyses in which an additional covariate was included for: body mass index (BMI), education, systolic blood pressure (SBP), total cholesterol (TC), self-reported alcohol consumption (alc), or a polyepigenetic risk score for alcohol consumption (PERS).

**Supplementary Table 12**. Differentially methylated positions (DMPs) associated (*P* ≤ 3.6 × 10^−8^) with the CAIDE2 dementia risk score in the discovery sample (n = 3076). For each DMP, the symbol of the gene in which the probe maps (if any), probe location (CHR and MAPINFO; hg19/GRCh37), location relative to genes (FEATURE; Body: within the gene body; TSS200: within 200 bp of the transcription start site; TSS1500: within 1500 bp of the transcription start site) and CpG islands (CpGISLAND), estimated effect (Beta), standard error of the estimated effect (Beta SE), t-statistic and *p*-value are shown.

**Supplementary Table 13**. Differentially methylated positions (DMPs) showing replicated association with the CAIDE2 dementia risk score. DMPs identified in the discovery sample (n = 3076; *P* ≤ 3.6 × 10^−8^) that showed association (*P* ≤ 0.05/18) with a consistent direction of effect in the replication sample (n = 2802) were deemed to have replicated. The probe ID, symbol of the gene in which the probe maps (if any), probe location (CHR and MAPINFO; hg19/GRCh37), genomic features associated with the probe (Body: within the gene body; TSS200: within 200 bp of the transcription start site; TSS1500: within 1500 bp of the transcription start site), beta coefficient, standard error of the beta, t-statistic and p-value in the discovery (dis) and replication (rep) samples are show for each replicated DMP.

**Supplementary Table 14**. Differentially methylated positions (DMPs) associated (*P* ≤ 3.6 × 10^−8^) with the CAIDE2 dementia risk score in an inverse standard error-weighted fixed effects meta-analysis of the discovery and replication samples (total n = 5878). For each DMP, the probe location (CHR and BP; hg19/GRCh37), symbol of the gene in which the probe maps (if any), direction of effect in the discovery and replication samples, estimated effect, standard error of the estimated effect (SE) and *p*-value are shown.

**Supplementary Table 15**. EWAS catalog traits/conditions showing significant enrichment for the CAIDE1-associated meta-DMPs. For each trait or condition, the number of DMP-harbouring genes that have been suggestively (*P*≤1 × 10-5) associated with the phenotype (sig_freq), the number of non-significant CpG-harbouring genes have been suggestively (*P*≤1 × 10-5) associated with the phenotype, and the significance (*P* value) and odds ratio (OR) of the enrichment are shown.

**Supplementary Table 16**. DMRs associated with the CAIDE1 risk score in a meta-analysis of the discovery and replication samples (total n = 5879). For each DMR, the location (Chr., Start and End, hg19/GRCh37), length, effect size (effect) and standard error of the effect (SE), Bonferroni-adjusted *p*-value (Adj. p-value), number of CpGs in the DMR (N), and the identities of these CpGs (CpGs) are shown.

